# A Methodological Framework for the Efficient Characterization of Peripheral Nerve Stimulation Parameters

**DOI:** 10.1101/2025.03.29.25324704

**Authors:** Rachel S. Jakes, Benjamin J. Alexander, Vlad I. Marcu, A. Bolu Ajiboye, Dustin J. Tyler

## Abstract

*Objective.* Restoring movement and somatosensation with peripheral nerve stimulation (PNS) requires precise neural activation. Pulse amplitude (PA) and pulse width (PW) modulate neural response differently, offering potential for improved selectivity. However, simultaneously modulating both parameters is rare due to the time required to map the two-dimensional space. This paper proposes and clinically validates an efficient method to characterize multiple intensities in the PA-PW space for motor and perceptual sensory applications. We also explore distinct activation patterns and potential applications of stimulation within the two-dimensional space. *Approach.* We used cuff electrodes implanted in one participant with a spinal cord injury to generate equal muscle activation contours and two participants with upper limb loss to generate perceptual intensity contours across multiple intensities in the PA-PW space. Strength-duration (SD) curves were mapped to the contours using varying sample point subsets and assessed for fit. Finite element modeling of the human nerve and activation simulations evaluated differences in recruited axon populations across the PA-PW space. *Main results.* SD curves accurately fit all levels of motor activation and perceptual sensory intensity (median *R*^2^ = 0.996 and 0.984, respectively). Reliable estimates of SD curves at any intensity require only two sufficiently spaced points. We propose a novel method for efficiently characterizing the PA-PW space utilizing the SD curve, including a metric to assess estimation accuracy based on the sampled points. *In silico*, intensity-matched high PW and high PA stimulation activated unique axon subsets, with high PA stimuli preferentially recruiting large-diameter motor and sensory axons farther from the contact. *Significance.* This work supports the use of SD curves to efficiently define a two-dimensional stimulation region for clinical motor and sensory PNS. Applying the proposed characterization method could enhance selectivity and resolution, reducing fatigue, improving fine motor control, and enabling unique percept generation. (ClinicalTrials.gov ID NCT03898804)

## Introduction

Peripheral nerve stimulation (PNS), or the use of electricity to activate peripheral nerves, is a powerful tool with the capacity for regulation and restoration of neurological function. For patients with spinal cord injuries or stroke, a type of PNS called functional electrical stimulation (FES) can circumvent the injuries in the central nervous system by functionally activating the peripheral motor neurons that innervate relevant muscles [1,2]. For people with limb loss, sensory PNS can activate the neurons that formerly innervated the missing limb, reestablishing the sense of touch for sensory neuroprostheses [3]. For these and other PNS applications, modulation of the magnitude of the nervous system’s output is vital. Dexterous motor control requires the ability to smoothly adjust the activation of multiple muscles, while the ability to distinguish between light touch and a firm grasp is critical for handling fragile objects or holding someone’s hand. The magnitude of the neural response at the level of muscle activation and somatosensory perception is a function of both how many axons are activated and how often those axons fire. The number of axons that fire is modulated by both the pulse amplitude (PA) and the pulse width (PW) of the electrical stimulus.

Modulation of both PA and PW in a two-dimensional space should enable better control of neural activation, as PA and PW have been shown to have differential effects on axon recruitment [4–6]. Historically, charge-modulated intensity has been mapped and modulated in one dimension, varying PA or PW and holding the other parameter constant. While this method does modulate intensity, its one-dimensional axis limits the order and magnitude of certain axon populations’ activation. Prior *in silico* work has shown that a high PA and low PW stimulation paradigm is more likely to recruit large diameter fibers at threshold than low PA and high PW [4,5]. Stimulation paradigms at high PA compared to high PW have also exhibited different degrees of selectivity as a function of distance from the stimulating electrode in *in silico* and in *in vivo* animal motor studies [5,6]. A clinical study showed differential activation of the Hoffman (H) reflex at different PA and PW paradigms, all intensity matched for equal M-wave recruitment [7]. Though there is evidence of systematic differences between high PA and high PW stimulation, these differences have rarely been functionally applied.

Iso-intensity differential axon activation via simultaneous PA and PW modulation has many potential clinical uses. For motor control, the ability to modulate the activation of different muscles independently by varying both PA and PW has the potential to significantly increase the dexterity and degrees of freedom of existing motor neuroprostheses. Intra-muscle selectivity, the ability to activate different motor units within the same muscle, has been demonstrated to be highly beneficial in minimizing fatigue [8–11]. By cycling between different groups of motor units within the same muscle, it is possible to maintain a constant force while spreading the load over a larger population of muscle fibers. Simultaneous parameter modulation could also enable shallower paths from threshold to maximum, which has long been hypothesized to enable more precise movements [12–15]. For sensory restoration, more specific control of axon population recruitment could enable the modulation of location or quality with a single contact, independent of intensity. For both motor FES and sensory PNS, multidimensional charge modulation may increase the usable range of intensity by identifying how pain or contraction thresholds change across the space. While the functional potential of a two-dimensional charge modulation space is high, the primary barrier to utilizing the full extent of this charge modulation space is the time to calibrate stimulation in both dimensions.

Despite the many potential benefits of modulating the PA-PW space when activating motor or sensory neurons, it is almost never done. Instead, typically, charge is modulated via either PA or PW control while holding the other parameter constant. For motor neuroprostheses, recruitment curves have been the field standard for decades [16–22]. However, each recruitment curve provides only a single slice of the complete stimulation-space, failing to capture the maximum selectivity of different motor units and muscles. For sensory neuroprostheses, while perceptual threshold is sometimes characterized in two dimensions [23–25], standard intensity modulation procedures typically choose either PW or PA modulation and keep the other constant, as defined by the charge term of the activation charge rate equation [26–29]. Pulse frequency (PF) is also sometimes used to modulate sensory intensity, but because PF impacts perceived intensity less than charge [26,28] and additionally alters sensation quality [27,30], frequency modulation is less suitable than charge modulation for independent intensity control.

In both sensory and motor neuroprostheses, a key reason for the lack of simultaneous PA-PW modulation is the prohibitive amount of time it would take to characterize the outputs of all possible parameter combinations. For motor FES, single-pulse twitch-activation can be used to approximate the motor output of continuous tetanic stimulation [31]. By measuring force [32] or electromyographic [19] twitch outputs it is then possible to quickly characterize muscle responses to electrical stimulation. However, for systems with many, multi-contact electrodes, even this methodology can quickly become prohibitively time consuming. A method for efficient characterization based on a 3D extrapolation of the Gompertz equation has been described, but it does not have a firm physiological basis and has not been widely used [33]. For sensory activation, the duration of stimulation-space mapping is an even greater challenge due to the required human-in-the-loop aspect of sensory percept characterization. As an added complication, single pulse and continuous stimulation trains have been shown to evoke different percepts, so stimulation trains must be used for functional percepts for sensory neuroprostheses [24]. For motor and sensory neuroprostheses to make use of the full PA-PW stimulation space, a faster method of characterization is necessary.

Our proposed method for efficient multi-dimensional stimulation space characterization is to decrease the necessary number of sampled points by applying the well-validated neural feature, the strength-duration (SD) curve (**Eq. 1**),

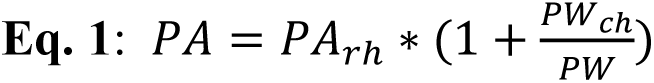

where PA is pulse amplitude, PW is pulse width, PA_rh_ is the rheobase, and PW_ch_ is the chronaxie. The equation describes the relationship between the PA (strength) and PW (duration) of a square wave required to create the same level of evoked neural activation. The rheobase is the amplitude required for activation at infinite PW, while the chronaxie is the PW on the curve when the PA is double the rheobase. The SD curve was first described as an axon-level characteristic of the intracellular threshold of activation by Weiss in 1901 [34]. Though other models have been proposed, the Weiss model (**Eq. 1**) has been shown to be the most accurate *in silico* and *in vivo* [35–37]. It has since been demonstrated that Weiss’ SD curve accurately scales from the single fiber response that it was initially defined for to a collective response at multiple levels of activation as quantified by compound muscle and compound sensory action potentials (CMAPs and CSAPs) [36,38]. In addition to the objective measurement of CSAPs, sensory perception induced by PNS has been shown to obey this same relationship at the threshold of activation [23,39,40]. However, it remains unknown if SD curves can accurately describe perceptual intensities at higher percept magnitudes across the dynamic intensity range.

Given the apparent robustness of the strength-duration relationship and the fact that the equation has only 2 parameters, it has been suggested that 10, 5, or even 2 points may be sufficient to accurately compute the entire curve [23,36]. Recent *in silico* work, based on the Weiss equation, suggested that the furthest possible two points on the curve could be the optimal samples to accurately construct the curve [41]. No modeling work involving axon models or neural channel dynamics have validated these results. Further, no *in vivo* research has investigated the ideal points to fit motor and somatosensory SD curves or how the accuracy of these fits varies with different parameter pairs or levels of activation.

Establishing the SD curve as a reliable tool for PNS parameter space characterization requires examining several key physiological and methodological factors. Firstly, the SD curve must be a consistently accurate representation of the system neural activation that we are interested in characterizing. We therefore demonstrate that the SD curve equation accurately fits evoked EMG and perceived intensities measured from human participants across the full functional range of activation. Secondly, optimizing SD curve sampling efficiency and accuracy is essential for its application in FES and sensory PNS activation. To address this, we identified the most effective points for constructing SD curves efficiently and assessed the robustness of curve generation when using non-optimal samples. Finally, since a major potential benefit of the combined PA and PW modulation is the ability to activate different axonal populations, we used *in silico* models of the human nerve to characterize the differences in the populations activated at high PA and high PW conditions while maintaining a similar number of activated fibers.

## Methods

### Clinical

#### Motor

##### Participants

One male in his thirties with a motor complete C4 level spinal cord injury participated in motor data collection as part of the “Reconnecting the Hand and Arm to the Brain (ReHAB)” clinical trial [42]. Eight composite flat interface nerve electrodes (C-FINEs) [43,44] were implanted around the participant’s median, radial, ulnar, musculocutaneous, lateral pectoral, and axillary, suprascapular, and long thoracic nerves 5 years post injury. Data collection for this paper started four years after implantation and consisted of six four-hour sessions that occurred over four-months. The U.S. Food and Drug Administration approved an Investigational Device Exemption (ClinicalTrials.gov ID: NCT03898804) for the use of the peripheral nerve electrodes in this study. The University Hospitals Cleveland Medical Center institutional review board approved the protocol and provided study oversight. The participant provided informed consent for the study.

##### Stimulation Delivery and Calibration

A Windows PC running a custom search method on MATLAB 2024a (*MathWorks, Natick, MA, US)* determined stimulation parameters and sent them to the stimulator through an intermediary xPC realtime target. Electrical stimulation was provided by a custom current-controlled universal external control unit (UECU) stimulator [45]. The PW resolution of the stimulator was 1 µs from 1 to 250 µs. The PA resolution of the stimulator was 0.1 mA below 2 mA and 1 mA at higher amplitudes. The stimulator delivered single pulses of a cathode-first charge-balanced biphasic square wave between one of the cathodic contacts and the anode strips on the C-FINE. We quantified muscle activation using 3 trial-averaged EMG responses rectified and integrated from 5 to 45 ms after twitch stimuli [19]. The Bluetooth enabled surface EMG sensors of the Trigno Wireless Biofeedback System (*Delsys, Natick, MA, USA*) recorded EMG. We tested two contacts on each of five different nerve cuffs by stimulating the contact and recording surface EMG from one large and functionally relevant muscle or muscle group innervated by the given nerve (Table 1).

**Table 1:** Motor Nerve Cuffs, Contacts, and Muscles Analyzed. Five nerve cuffs, each on a different nerve in the arm or shoulder, were tested for the acquisition of muscle activation data. For each nerve cuff, one contact on each side of the cuff was stimulated and one large muscle or muscle group that the nerve is known to innervate was recorded through surface EMG.

| **Nerve Cuff** | Axillary | Lateral Pectoral | Musculocutaneous | Median | Radial |
| --- | --- | --- | --- | --- | --- |
| **Contacts Stimulated** | 4,13 | 5,15 | 6,13 | 4,9 | 5,12 |
| **Recorded Muscle or Muscle Group** | Posterior Deltoid | Upper Pectoralis | Biceps | Wrist and Finger Extrinsic Flexors | Wrist and Finger Extrinsic Extensors |

##### Equal Muscle Activation Contour Generation

Equal muscle activation contours map the relationship between two variables and their relative effect on the resulting muscle activation. For each contour, a specific target EMG output was defined and our experimental setup (**Figure 1**) used binary search to acquire the set of PA-PW pairs that resulted in the EMG values closest to that target.

**Figure 1:**
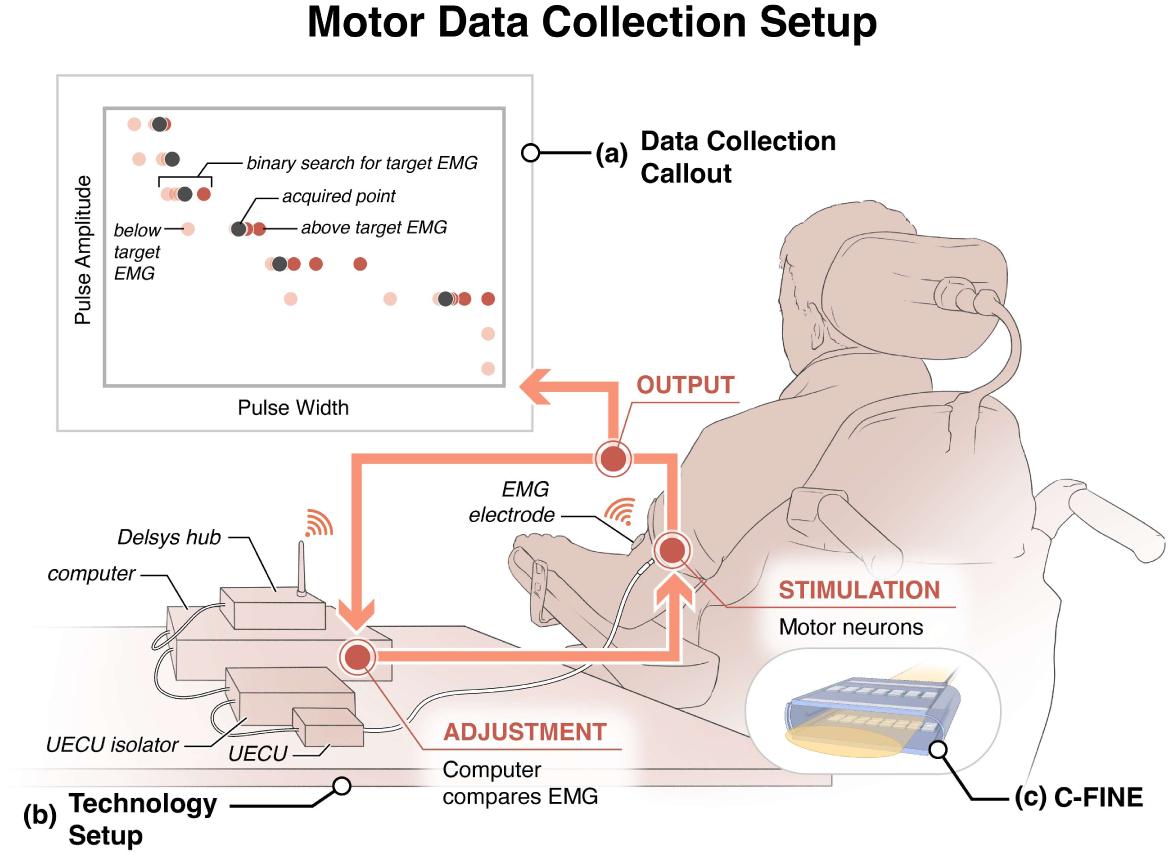
Experimental Setup for Muscle Activation Contour Collection. **(a)** Equal intensity muscle activation contours were acquired using a binary search algorithm that determined the optimal stimulation parameters to match a target EMG level. The binary search method started at a pulse amplitude (PA) of 0.1 mA and pulse width (PW) of 250 µs. The computer increased the PA until the recorded EMG value was above the target EMG level. At this point, the computer performed binary search to find the PW that provided the closest EMG value to the target. A repeated binary search at increasing amplitudes through all possible PAs completed the contour. **(b)** The UECU stimulator provided electrical stimulation to the nerve cuff electrodes in the participant’s arm. A Delsys Trigno EMG sensor recorded the EMG voltage which the computer then processed. **(c)** Stimulation was delivered through C-FINE cuffs containing 15 cathodic contacts and an anode strip.

We acquired equal muscle activation contours for 10, 30, 50, 70, and 90 percent of the maximum evoked muscle activation for 10 different muscle-contact pairs. This maximum was established prior to data collection by finding the upper asymptote of a pulse width modulated recruitment curve. Our binary search method acquired each contour (**Figure 1**) by starting at 0.1 mA and 250 µs and increasing PA in steps of 0.1 mA until it recorded an EMG value above the target threshold. At this point the method varied PW from 1 to 250 µs using binary search until it determined the PW value that resulted in muscle activation closest to the target level. The sampling method then moved up to the next PA value, using the final PW value from the previous PA as the new maximum stimulus value. We updated the maximum allowable PW in this way to prevent painful, high intensity, stimuli and with the assumption that increased PA, and identical PW should always produce the same or greater levels of muscle activation. The system continued this process until the maximum PA value of 3.0 mA was reached. We acquired 3 blocks each consisting of all 5 activation levels with their acquisition order randomized within each block to remove any dependence on order.

#### Sensory

##### Participants

Two participants (both male) with upper limb loss participated in sensory data collection. Subject 1 received a left transradial amputation from traumatic injury 11 years prior to this study and was implanted with two 16-channel C-FINEs around the median and ulnar nerves in the residuum 3 years later, 8 years prior to data collection for this study. Subject 2 received a right transhumeral amputation from a traumatic injury also 11 years prior to this study and was implanted with two 16-channel C-FINEs around the median and radial nerves in the ipsilateral axilla 7 months prior to data collection for this study. The sensory data in this paper comprises one four-hour session with Subject 1 and three four-hour sessions with Subject 2 over four months, all sampled from the median cuff. Median nerve stimulation could elicit both motor and sensory percepts in Subject 1 but only sensory percepts in Subject 2 due to the absence of the corresponding innervated muscles. The U.S. Food and Drug Administration Investigational Device Exemption, the Cleveland Department of Veterans Affairs Medical Center Institutional Review Board, and the Department of the Navy Human Research Protection Program approved all study devices and procedures. All participants provided informed consent for the study.

##### Stimulation Delivery and Calibration

All stimuli were 100 Hz trains of biphasic cathode-first charge-balanced square wave pulses between one of 15 cathodic contacts and an anode strip on the median nerve C-FINE. Stimulation parameters set in MATLAB R2020a (*MathWorks, Natick, MA, USA*) drove a current-controlled Grapevine Neural Interface Processor (*Ripple Neuro, Salt Lake City, UT, USA*).

To assess each participant’s approximate perceptual range at a long PW (250 µs), we applied an increasing PA staircase with 2 reversals and a step size of 0.01 mA. Participants assigned the perceptual threshold and upper bound intensity numbers via the magnitude estimation methodology described by Graczyk et al. [26]. The upper bound was defined as the first of either maximum comfortable sensation or a muscle contraction. Participants also identified perceptual threshold and maximum comfortable intensity via an increasing PW staircase at a PA of 2.0 mA. Last, participants labeled PAs at a PW of 250 µs that produced sensations at about 25%, 50%, and 75% of their dynamic intensity range as defined by their minimum and maximum magnitude estimations. These five PAs across the dynamic intensity range became the reference values for equal intensity contour generation. Over the course of the session, the estimated magnitude of these reference stimuli sometimes shifted, but the stimulation parameters remained constant as long as the participant still reported the stimuli as perceptible and comfortable.

##### Equal Somatosensory Perceptual Intensity Contour Generation

Equal intensity contours map the relationship between two variables and their relative effect on perceived intensity. In haptic feedback research, equal intensity contours have been generated between frequency and amplitude of a signal using the method of adjustment [46,47]. Our experimental design (**Figure 2**) modifies those methods by determining the PWs at which two different PAs of stimulation feel equal in intensity.

**Figure 2:**
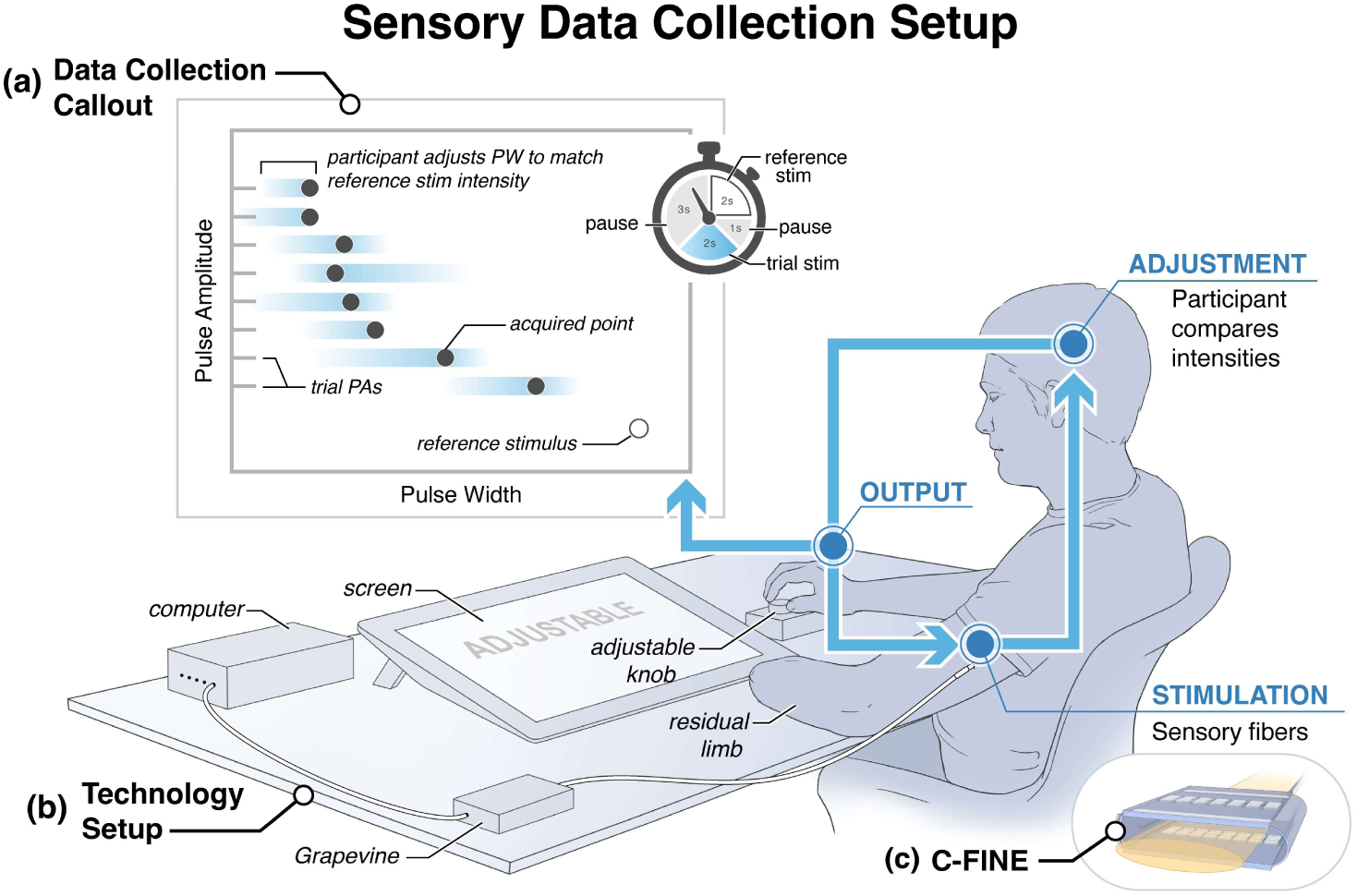
Experimental Setup for Sensory Perceptual Intensity Contour Collection. **(a)** Equal intensity contours were collected using a method of adjustment, in which participants used an adjustable knob attached to a rotary encoder to adjust pulse width at set pulse amplitudes to match the intensity of a reference stimulus. Reference and trial stimuli were alternated as shown in the timer icon. **(b)** The Ripple Grapevine provided stimulation through percutaneous wires in the participant’s residual limb to a cuff electrode around the participant’s median nerve. **(c)** Stimulation was delivered through C-FINE cuffs containing 15 cathodic contacts and an anode strip.

We defined a single intensity contour as nine points: the reference PA and eight trial PAs sampled above the highest reference PA for the contact. One intensity contour was created per reference stimulus per block. To generate an equal intensity point, participants received two seconds of a reference stimulus, i.e., a reference PA at 250 µs PW, followed by two seconds of a trial stimulus, defined as a trial PA and an adjustable PW. A 1 second and 3 second pause followed the reference and trial stimuli, respectively, to minimize adaptation, as recommended by Van Doren [46]. Participants used a rotary encoder knob to adjust the PW of the trial stimulus. The participant could turn the PW knob at any point during the trial and the trial PW would update to the new value the next time the trial stimulus was provided. This pair of stimuli repeated until the participant felt the reference and trial stimuli were identical in intensity. To account for anchoring bias, the adjustable PW started at or below perceptual threshold for each trial. We also placed limitations on the adjustable stimulation that ensured participant safety while still providing sufficient modulation space for participants to place the trial stimulus anywhere in their dynamic range.

Each equal intensity contour was collected in a randomized order over three one-hour blocks for a total of 27 points per contour. In some cases, full contours could not be collected for weaker intensities due to adaptation in the later blocks. Contours reported in this manuscript had a minimum of 3 trials per reference and trial stimulus pair and a minimum of 9 points per contour per block. In all, three full contours were collected from contact 2 on Subject 1, four contours were collected from contacts 1 and 6 on Subject 2, and five contours were collected from contact 5 on Subject 2. Some contours from contact 5 on Subject 2 had 4 or 5 trials per reference and trial stimulus pair, as repeat points were collected in the final block to better investigate any temporal effects.

#### Data Analysis

For both motor and sensory equal intensity contours, we first assessed if there was any significant difference across points accumulated in different blocks for the same equal intensity contour to determine if points should be grouped or separated by block for curve fitting. Two non-linear mixed effects models fit with the Weiss equation (**Eq. 1**) were applied to each equal intensity contour, one accounting for block number as a random effect and one without random effects. The fits were compared with a likelihood ratio test. If block number had a significant effect on the model, curve fits would be completed individually for each block. Otherwise, blocks would be grouped together for curve fits. In accordance with this methodology, motor contours from different blocks were treated as unique contours, while sensory contours were combined across blocks and treated as samples of the same contour.

To assess the ability of the Weiss SD curve to describe the PA-PW equal intensity contours, we applied **Eq. 1** to each contour using a non-linear model with an iterative least squares estimation fit algorithm. Comparing the resulting curve to the points in the equal intensity contour yielded a goodness-of-fit R^2^ value.

To evaluate if the SD curve solved as a system of equations with just two (*PW*, *PA*) points could accurately describe an equal intensity contour, an end-points method and any two-point fit method was used. Previous mathematical modeling suggests that the points sampled furthest apart in the PA-PW space, one each in the vertical and horizontal regions of the curve (i.e., the ‘end-points’), have the highest probability of returning a good fit (37). The any two-point method was used as a comparison because the position of the ‘end-points’ is somewhat arbitrary. In the motor analysis, no trial averaging was done across contours, while in the sensory analysis, both methods were tested with and without trial averaging of the solver points. In all cases the resulting models were compared back to the sampled equal intensity contour via an R^2^ goodness-of-fit calculation.

A Kolmogorov-Smirnov test assessed the normality of each R^2^ distribution. As the distributions were non-normal, a Kruskal-Wallis test compared R^2^ values across three motor and five sensory distributions to test for differences in goodness of fit based on the number and selection of points used to create the SD curve. Non-parametric Dunn-Šidák tests with multiple comparison corrections followed.

Last, to identify the threshold at which fit accuracy is lost based on selection of solver points, we plotted goodness of fit as a function of the ratio of the slopes, or slope ratio (SR) of the SD curve at the two solver points for both the motor and sensory data sets (**Eq. 2**). Slopes were evaluated as the derivative of the full fit SD curve (**Eq. 3**) at the PA of each solver point, in which *PA_i_* < *PA_j_*.

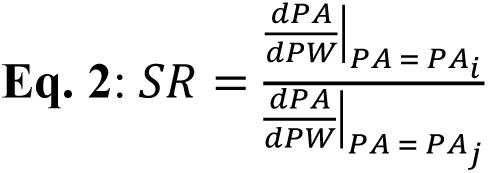

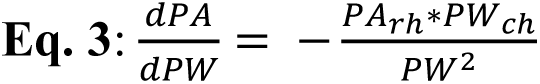

Because of the positive concavity of the SD curve, SR will always be between 0 and 1, with further apart points yielding an SR closer to 0. SR is also unitless so it can be standardized across paradigms and sample spaces.

#### Modeling

We used simulations of peripheral nerve stimulation to examine differences in axonal activation at high PA and high PW stimulation at equal intensities of activation. Finite element models based on the histology of human peripheral nerve samples provided estimations of axon population activation. Electrode configurations based on 16-contact C-FINEs [43,44] were placed in the model. The conductivities of the different neural tissues were assigned in accordance with empirical data on the resistivities of the endoneurium [48], perineurium [49], and epineurium [50], as well as saline [51]. To determine voltage along different axons, a 1 mA current was simulated at each of the 16 contacts, and the resulting voltage fields were recorded within each fascicle. All simulated stimuli were biphasic square currents with a 2:1 charge recovery ratio and no inter-pulse interval.

Simulating the axonal population response to a given monopolar stimulus was done in MATLAB 2024a (*MathWorks, Natick, MA, USA*) by solving the Gaines cable model [52] in response to externally applied voltage fields as solved for in COMSOL (*COMSOL, Burlington, MA, USA*). The Gaines model captures differences in ion channel densities between motor and sensory axons, enabling the identification of distinct axonal dynamics. Each simulated fascicle contained 700 axons with randomized internode offsets, positions, and physiologically informed diameters based on sensory and motor populations. To vary the stimulation PA, we linearly scaled the COMSOL voltage fields from the 1 mA template. This PA modulation method aligns with previous studies that suggest peripheral neural tissue is electrically linear with minimal voltage-dependent impedance changes [53–55]. We simulated axonal responses to different PWs by varying the duration of external voltage application in the Gaines model.

The simulation model distinguishes sensory and motor axons by assigning each axon population distinct channel conductances [52] and axon diameter distributions. We assign sensory axons diameters based on the distribution of the human sural nerve [56], as it is a primarily sensory nerve. The motor axon diameter distribution was obtained by starting with the human median nerve axon distribution [57] and proportionally subtracting the sural nerve distribution, thus isolating only motor axon diameters.

Multiple SD curves were generated by simulating monopolar stimulation along all 15 stimulating contacts of the simulated C-FINE at nine pulse amplitudes between 0.1 and 0.6 mA and ten pulse widths between 10 and 250 µs. We generated an exhaustive set of PA and PW pairs and recorded their corresponding neural activations. We then grouped stimuli into ‘iso-intensities,’ with each stimulus in an iso-intensity activating within 1% of the same total axon count. We classified stimuli as in the vertical region of the SD curve if their pulse widths were 50 µs or less and as in the horizontal region of the curve if they were 100 µs or more. We compared the resulting iso-intensities to the SD curve (**Eq. 1**) by calculating R² values of the Weiss equation fits to iso-intensities in both the motor and sensory nerve.

Using the simulated data, we measured the axon diameters uniquely activated between stimulation in the vertical and horizontal regions of the SD curve. We also quantified the distances from activated axons to the stimulating contacts under these conditions. T-tests were used to determine if axon diameters and average axon-to-contact distances differ significantly between the horizontal and vertical regions of the same SD curve. Finally, we compared the differences across SD curves that activate various percentages of the model’s axons and assess whether low-intensity stimulation exhibits trends distinct from high-intensity stimulation.

## Results

### Clinical

#### SD curves describe equal muscle activation and perceptual intensity contours at high goodness of fit

The Weiss SD curve equation was found to describe equal muscle activation and perceptual intensity contours across the respective activation ranges, with median R^2^ values of 0.996 and 0.984, respectively (**Figure 3**).

**Figure 3:**
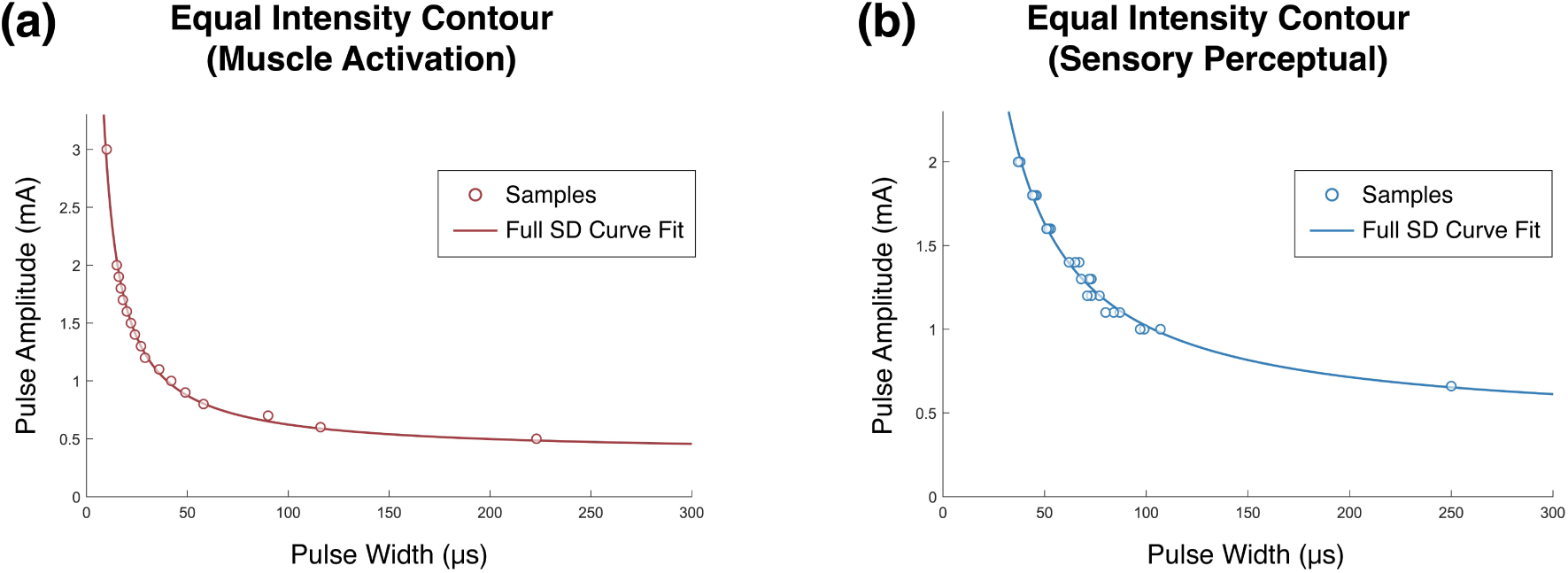
Equal Intensity Contours with SD Curve Fits. Data points collected for the equal intensity contours are shown in circles, while the Weiss SD curve (Eq. 1) fit to those data is a solid line. **(a)** This contour was collected for 10% EMG activation of the biceps muscle of the motor participant. A total of 17 points at unique PAs were collected from a single block. The given SD curve fit has an R^2^ value of 0.995. **(b)** This contour was collected for a mid-intensity sensory percept from Subject 2, with a reference stimulus amplitude of 0.66 mA. Points at nine PAs were collected in three blocks for a total of 27 points. The given SD curve fit has an R^2^ value of 0.990.

#### Two Points can accurately solve for SD curves at multiple levels of neural activation

The two unknowns of an SD curve, rheobase and chronaxie, can be solved for as a system of equations by providing two points. These 2-point solutions were solved using the points furthest into the horizontal and vertical regions of the SD curve and compared to the fit with the full data set (**Figure 4**). In the case of the sensory data (**Figure 4(b)**), because three samples were collected for the highest point in the vertical region, the average of those three values was used to solve the system of equations.

**Figure 4:**
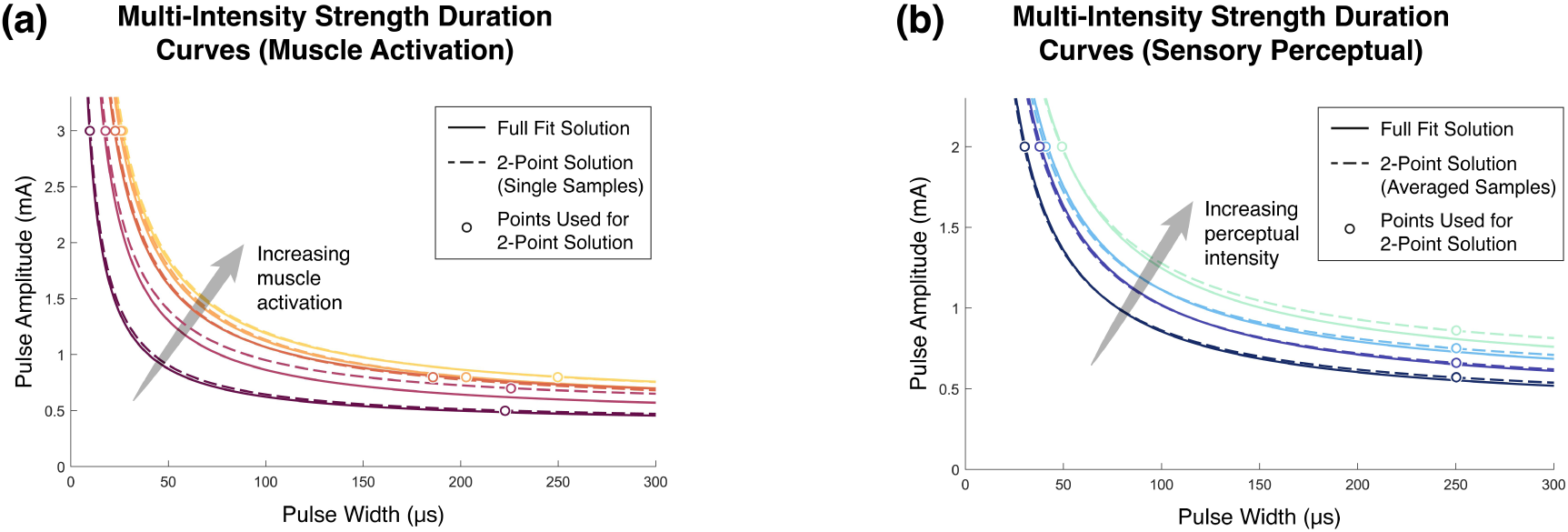
Multi-Intensity Fit and Solved SD Curves. SD curves that were fit with a full contour, as in **Figure 3**, are shown with solid lines, while SD curves that were generated by solving for the Weiss equation with the two points that are furthest into the horizontal and vertical regions are shown with dashed lines. Lines of the same color correspond to the same contour. **(a)** These curves are generated from activation of the biceps muscle, with each contour representing 10%, 30%, 50%, 70%, or 90% of the asymptotic maximum of that muscle. **(b)** These curves are generated from activation of sensory fibers in the median nerve of Subject 2 that primarily result in percepts on the thumb and palm of the phantom limb. The curves shown span approximately 20% to 80% of the participant’s dynamic intensity range.

To assess how robust the quality of the solved SD curve is to point selection and trial-by-trial variance, curves were fit with all contour points, the two end-points, and every combination of two points in the contour. Due to small but significant differences between blocks for motor data, all motor SD curves were fit and evaluated with data from a single block (**Figure 5(a)**). For sensory data, the two-point solved curves were calculated with both trial-averaged points and single-trial points and evaluated against data from all blocks for the given contour (**Figure 5(b)**).

**Figure 5:**
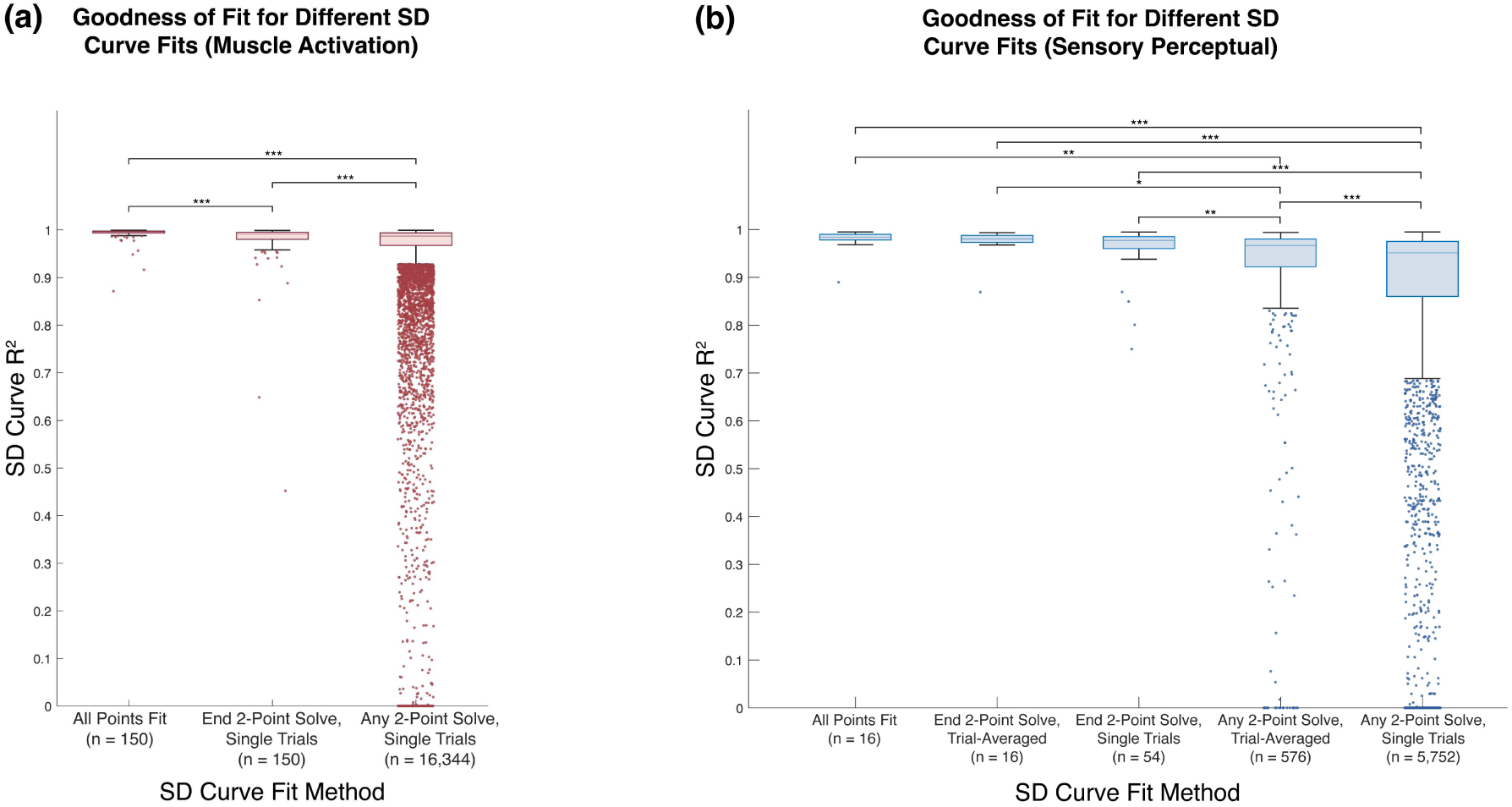
Distribution of R^2^ Values by SD Curve Fit Method. Outlying values, defined as values more than 1.5*IQR away from the 25^th^ or 75^th^ percentile, are jittered points below the boxes. Outliers below zero were set to 0 for visualization. Two-point pairs with no solution were set to an R^2^ of 0. Significant differences between groups as determined by a Dunn-Sidak post-hoc multiple comparison are shown with asterisks above pairings (*\* p<0.05, ** p<0.01, ***p<0.001*). **(a)** The muscle activation SD curves fit with full dataset, solved with end-points, and solved with any two points have median R^2^ values of 0.996, 0.991, and 0.987 respectively. Outliers represent 10%, 9.3%, and 14.0% of each dataset respectively. **(b)** The sensory perceptual SD curves fit with the full multi-block dataset, solved with trial-averaged end-points, solved with single-trial end-points, solved with any two trial-averaged points, and solved with any two single-trial points have median R^2^ values of 0.984, 0.980, 0.977, 0.967, 0.951, respectively. Outliers represent 6.3%, 6.3%, 7.4%, 14.6%, and 14.9% of each dataset, respectively.

Because of the wide differences in group size and non-normal distribution of the R^2^ values, a Kruskal-Wallis test compared the goodness of fits of the groups. The Kruskal-Wallis test showed significant differences for both motor and sensory data sets (both *p* < 0.001). Dunn-Sidak post-hoc tests showed significant differences between the distribution of R^2^ values for all SD curve solution methods for motor (all *p* < 0.001). For sensory SD curves, significant differences were found between the fits using any two single-trial points and every other methodology (*p* < 0.001). Differences with varying significance were also found between the solution with any two trial-averaged points and the other four methodologies. No significant difference was found between the goodness of fits of the SD curves fit with all points and the curves solved with trial-averaged or single-trial end-points.

In comparing the R^2^ distributions of the single-trial solutions, the interquartile ranges of the any-point solutions are about two and four times wider than those of the end-point solutions for muscle activation and sensory percepts, respectively. To further investigate the relationship between point selection and solution fit, R^2^ values are plotted as a function of the slope ratio (SR) (**Eq. 2**; **Figure 6**). Points sampled further apart on the curve, such as end-points, have an SR closer to 0, while points sampled closer together have an SR closer to 1.

**Figure 6:**
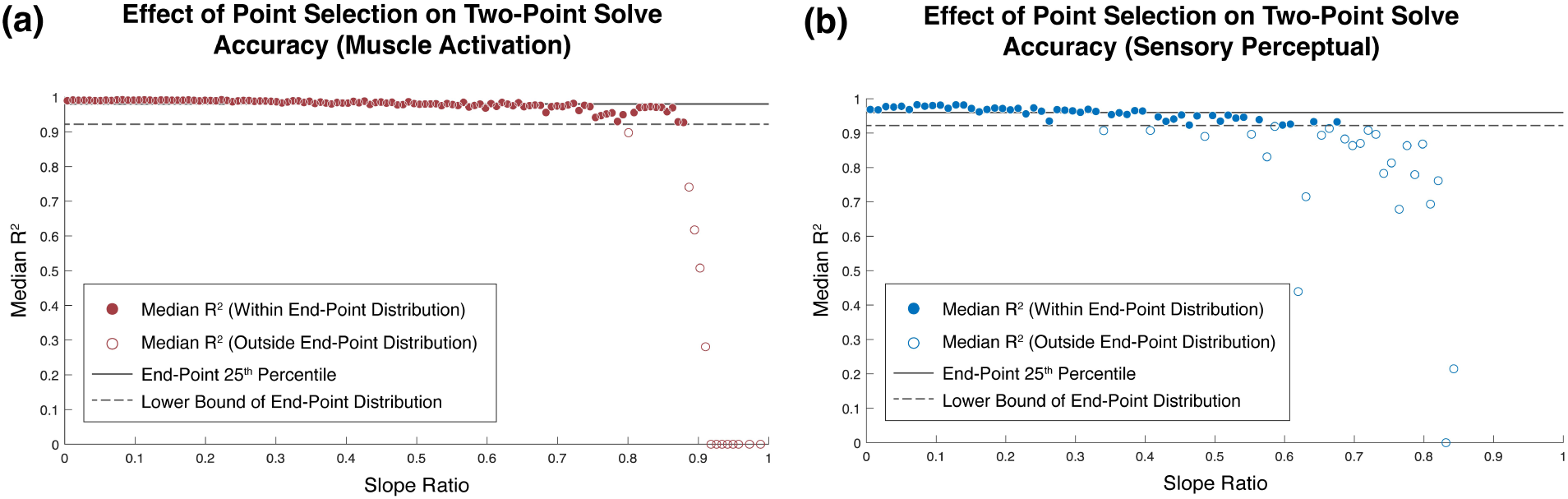
Two-Point R^2^ Fits by Slope Ratio at Solver Points. Point pairs with similar derivatives have a slope ratio closer to 1. Points were binned in the number of groups equal to the square root of the number of data points, with the width of each bin equal to the range of slope ratios divided by the number of bins. The y-coordinate is the median R^2^ value in the bin. The solid and dashed lines indicate the R^2^ values at the 25^th^ percentile and whisker of the single-trial end-point R^2^ distributions **(Figure 5). (a)** The motor data was binned into 128 groups, with a bin width of 0.0078 in slope ratio. End-point 25^th^ percentile and distribution boundary R^2^ were 0.980 and 0.958, respectively. (b) The sensory data was binned into 76 groups, with a bin width of 0.0111 slope ratio. The maximum slope ratio in the sensory data was 0.843 rather than 1 because the PA-PW space was sampled at a lower density. End-point 25^th^ percentile and distribution boundary R^2^ were 0.960 and 0.922, respectively.

To determine how far apart solver points must be on an SD curve to be classified as ‘end-points’, the any-point data was compared to the end-point R^2^ distribution. Lines indicate the 25^th^ percentile and lower bound of the single-trial end-point solution distributions. Points above these lines fall within the expected distribution of end-point goodness of fit. Point pairs with *SR* < 0.68 and *SR* < 0.34 fall within the end-point R^2^ distributions for muscle activation (**Figure 6(a)**) and sensory perception (**Figure 6(b)**), respectively. The highest SRs with a median goodness of fit within the end-point R^2^ distribution were 0.86 and 0.68 for motor and somatosensation.

#### Perceptual strength-duration curves do not significantly change across time despite adaptation in magnitude estimation

For all 16 sensory iso-intensity contours, block number did not significantly affect the non-linear mixed effects model (**Figure 7(b)**). This comparison includes the contours from the dataset collected across a mix of two standard 45-minute blocks and one repetitive block in which each reference and trial stimulus pair point was collected three times back-to-back. Despite the absence of significant difference found in the SD curves for each intensity across blocks, the participants rated 14 out of the 16 contours as changing in perceptual intensity across at least one of the blocks, ranging up to 50% of the magnitude estimation scale. Subject 1 reported the lowest intensity contour as changing across blocks but reported no difference in perceived magnitude for the higher intensities. Subject 2 reported a change in magnitude for all intensities and contacts (**Figure 7(a)**).

**Figure 7:**
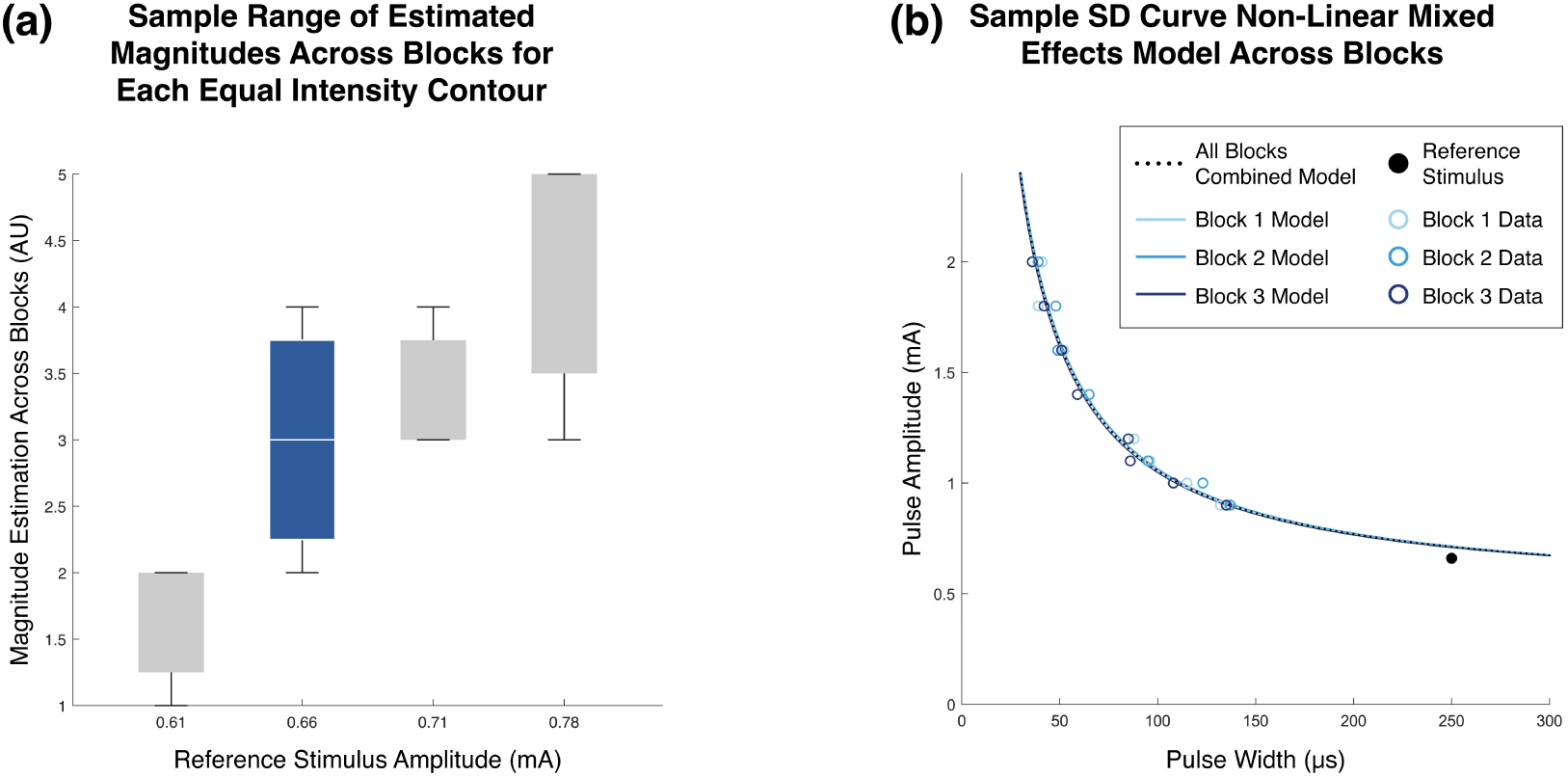
Changes in Perceived Sensation Magnitude and SD Curve Fits Across Blocks. Data is from stimulation on contact 1 of Subject 2, which elicits sensation on the middle finger of the phantom limb. **(a)** Each box displays the range of magnitudes assigned to each equal intensity contour, denoted by the reference stimulus amplitude given on the x-axis, across three blocks. Each reference stimulus elicited a distinct perceptual magnitude during calibration but may have shifted to alternative magnitudes by the beginning of block 1 due to adaptation during the calibration phase. **(b)** The data and SD curves for the box in blue in subfigure (a) are shown here. Each solid-line single-block model was generated using a non-linear mixed effects model with the block number as the random effect. The dotted black line is the non-linear model calculated without accounting for differences in blocks. The curves generated from blocks 1, 2, and 3 were rated as having magnitudes of 4, 3, and 2, respectively. No significant difference was found between the within-block curves despite the difference in reported magnitudes.

#### Different levels of muscle activation show no significant difference in R^2^ values for full and end-point fits

For each fit method used with the muscle activation data, the R^2^ values across the 10 muscle-contact pairs were binned by the percent level of activation (10%, 30%, 50%, 70%, 90%) of the given muscle. No statistically significant differences were seen for the full fit or the end-point fits. The full fit R^2^ values for the 5 levels of activation ranged from 0.994 to 0.997 while the end-point fit R^2^ values ranged from 0.988 to 0.992. However, the any two-point fit R^2^ values showed many significant differences, with the 90% condition being different from all other conditions (*p* < 0.01). R^2^ values toward the middle of the activation range tended to be higher than the R^2^ values closer to 10 and 90 percent. Despite this trend, the median R^2^ values still had a narrow range of 0.983 to 0.989.

### Modeling

We treated each activation level as a separate equal intensity contour and fit the corresponding recruitment surfaces to the Weiss equation to calculate an R² for each curve. Pooling these R² values for the 99 motor SD curves with more than two (*PW*, *PA*) points across activation intensities in the motor model produced a median value of 0.9995 (IQR 0.9981, 1); the sensory model yielded a similar median of 0.9981 (IQR 0.9927, 0.9993) from 91 simulated SD curves.

Across iso-intensities, two-sample t-tests show that the axon populations uniquely activated by high PA and high PW stimulation differ significantly (n ≥ 30) in both diameter and distance to the stimulating contact. High PW stimulation consistently activates axons closer to the simulated stimulating contact in both motor and sensory models (**Figures 8(a), 8(c)**), with 91.3% of sampled intensity pairs showing a significant difference in uniquely activated axon distance (88.4% *p* < 0.01, 2.9% *p* < 0.05). Of the significantly different pairs, only one showed high PA as activating axons closer to the contact. Similarly, high PW stimulation activates smaller axons than high PA stimulation across 95.7% of iso-intensity pairs (**Figures 8(b), 8(d)**) (94.2% *p* < 0.01, 1.4% *p* < 0.05). Overall, these results confirm that the mean axon diameters and distances to the stimulating electrodes of the neurons uniquely activated by paradigms on opposing ends of the SD curve differ significantly between the two stimulation methods.

**Figure 8:**
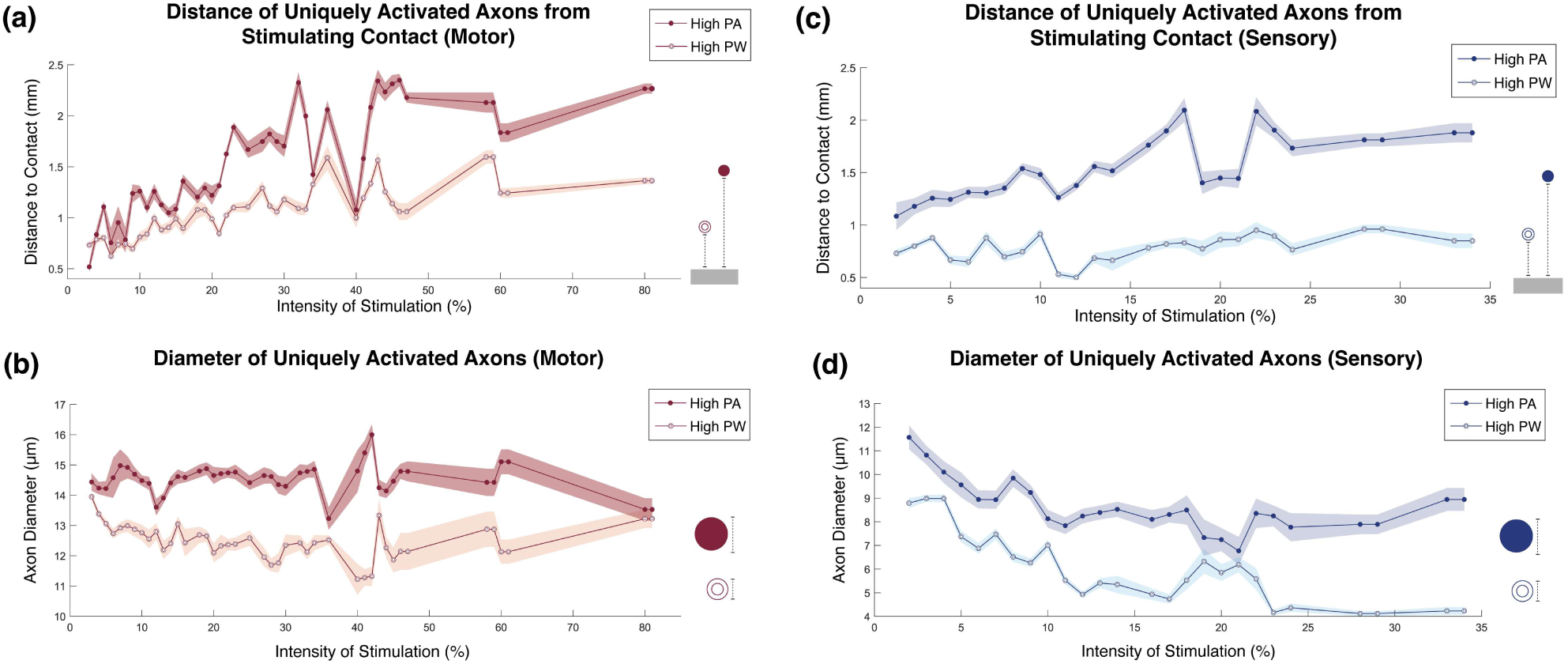
Axons Uniquely Activated by Intensity-Matched High PA and High PW Stimulation. Axons that are uniquely activated by either high PA (solid marker) or high PW (open marker) stimulation represent the expected differences in activation on the vertical and horizontal ends of an SD curve. Subfigures **(a)** and **(c)** show that high PW stimulation consistently activates axons closer to the cathodic contact compared to high PA stimulation, whereas **(b)** and **(d)** demonstrate that high PW stimulation uniquely activates smaller-diameter fibers comparatively. Vertical shaded regions represent 95% confidence intervals, with n ≥ 30 for each point. Motor models are in red, and sensory models are in blue.

While there are systematic differences in the axons uniquely activated between high PA and high PW stimuli at the same iso-intensity, the two stimulation paradigms share many activated axons. We applied the Dice similarity coefficient (DSC) to characterize the similarity between the axons activated by the high PA and high PW iso-intensity stimuli. A DSC of 0 indicates no shared axons, while 1 indicates identical populations. DSC values generally rose with increasing intensity, and a Kruskal-Wallis test followed by Dunn-Šidák post-hoc comparisons revealed that the lowest-intensity data differed significantly from high-intensity data in both motor and sensory populations. For both motor and sensory axon models, the DSC below 5% intensity was about 0.75, while at higher intensities the DSC leveled off to around 0.95.

## Discussion

### Accurate mapping of full functional ranges of muscle activation and perceived somatosensory intensity via SD curves

All clinically assessed intensities for both motor and perceived sensory neural activation fit the Weiss strength-duration curve accurately. For muscle activation, these results provide clinical validation of the SD curve in a person with a spinal cord injury, adding to previously established literature in healthy populations [36,38]. Further, the present study explores the accuracy of SD curves to describe muscle activation at levels above 60%, which is novel for all subject populations. The mapping of high-intensity muscle activation space is important for informing and improving the creation of sigmoidal recruitment curves and surfaces, which have dynamic curvatures at high intensities. The absence of a statistical difference between accuracy of fits across intensities for both full fits and end-point fits validates the use of the SD curve to accelerate the process of generating full recruitment surfaces.

For perceived somatosensory activation, the application of an SD curve to suprathreshold percept intensities is novel. While SD curves have been shown to hold for suprathreshold CSAPs in healthy populations [36,38], the human-in-the-loop aspect of determining multiple levels of perceptually equivalent neural activation adds a novel layer to generating the curves. Further, because Subject 2 does not have any muscles innervated by the median nerve in his residual limb, curves were able to be acquired at higher percept intensities without interference from muscle activation than might be possible in other participant populations. The expansion of perceptual SD curves beyond threshold also enables their use for exploring the PA-PW space for functional sensory restoration beyond baseline calibration and nerve health assessment [23,25,39]. Because the perceived magnitude of most curves varied across a session (**Figure 7(b)**), we did not describe or compare the acquired SD curves as a percentage of the perceptual dynamic range. Regardless, the data showed a consistently high accuracy of SD curve fits at all tested perceptual intensities (**Figure 5(b)**), validating their use to map the extrema of the perceptual intensity range.

### Accurate estimation of SD curves in as few as two points

The usefulness of the SD curve for describing the PA-PW space for clinical neural response modulation would be limited if the curve required many sample points to be accurately defined. Because of the human response component of sensory percept characterization, each single-block nine-point sensory curve took about 10 minutes to acquire, and it took three hours to fully characterize four percept intensity curves. While motor data collection was more efficient than sensory due to the automated stimulation adjustments, the average of 3 minutes to describe a single curve and 16 minutes to map five curves for a single contact pair is still too inefficient for functional parameter space characterization.

There was no statistical difference between the goodness of fit of a sensory percept SD curve fit with 27 sensory data points and one solved with two distanced points, either single trial or trial-averaged. For muscle activation, there were statistical differences, but the median R^2^ differed by only 0.005 between full and two-point motor SD curve fits. These results validate the use of two points to efficiently generate SD curves. This builds on Mogyoros’ clinical two-point SD curve model by showing that the accuracy of two-point solutions holds across all activation magnitudes and for perceptual intensity as well as compound action potentials [36]. Characterizing SD curves with only two points drastically reduces the time required to define them, increasing their functional applicability for mapping the full PA-PW space.

### Dependency of two-point SD curve accuracy on point sample position

In alignment with Alavi’s simulation-based findings [41], two-point fits were statistically significantly superior when sampled at the extrema of the parameter space in both motor and sensory activation (**Figure 5**). These results show that point position influences accuracy in clinical SD curve fits, and that there is a need for a method to assess if samples are close enough to the curve extrema to produce a quality SD curve.

To generalize point position independent of stimulator range or tissue impedance, the any-point curve fit distribution was expressed in terms of the ratio of the SD curve derivative at the points selected (**Figure 6**, **Eq. 2**). As expected, point pairs with more disparate derivatives, i.e. positioned on opposite sides of the elbow of the SD curve, produce better fits than pairs with similar derivatives. While the R^2^ fall-off is a continuum, cut-off values were based on the 25^th^ percentile and lower bound of the end-point R^2^ distributions for each respective modality. The any-point fits differ from the end-point distribution above slope ratios of 0.68 and 0.34 for motor and perceptual sensory curves, respectively. For the sensory distribution (**Figure 6(b)**), there are median R^2^ values within the end-point distribution up to a slope ratio of 0.68, but they are interspersed with lower R^2^ solutions, indicating a higher risk of a poor fit if the points are sampled in that space. Overall, a pair of points are likely to produce an accurate SD curve if the ratio of their slopes is less than 0.34 for perceptual sensory applications or 0.68 for motor applications. This analysis demonstrating where to sample SD curves for an accurate fit is novel in clinical literature.

With respect to whether trial-averaging the point pair is needed to generate an accurate SD curve, Forst et al. found that SD curve fit error tended to decrease with three to four trials per point in sensory surface stimulation [23]. However, these results did not consider the position of the sampled points. For the perceptual sensory SD curve fits, there was a significant difference between the trial-averaged and single-trial distributions when any two points were sampled but not when end-points were sampled (**Figure 5(b)**). Further, when expressed in terms of slope ratio, the trial-averaged any point distribution matched the single-trial distribution at low slope ratios, with the same R^2^ fall-off at 0.34. However, trial-averaged samples are more robust at higher slope ratios; while the last R^2^ value within the end-point distribution occurs at a slope ratio of 0.68 for single-trial point pairs, there are trial-averaged point pairs with R^2^ values within the end-point distribution up to 0.81. Overall, if the sampled point pair have a slope ratio less than 0.34, trial-averaging is not a necessary step to ensure a quality sensory SD curve fit. Otherwise, trial-averaging may have a positive impact on fit quality when sample points are closer together.

### Summary: Recommendation for how to efficiently collect SD curves

To most efficiently and accurately acquire an SD curve, we recommend sampling two iso-intensity points, one each at the highest possible PA and the highest possible PW allowed by the interface and stimulator. With respect to sensory sampling, ask participants to compare the intensity of the two points rather than relying on magnitude estimation to maximize the precision of the curve.

After collecting the two equal intensity points, the point pair should be used to solve the Weiss equation (**Eq. 1**). To ensure the collected points are far enough apart on the curve for an accurate estimate, calculate the derivative of the resulting SD curve at the location of the two sample points (**Eq. 3**). Calculate the slope ratio between the two points (**Eq. 2**), and check that the SR is less than 0.34 for sensory curve formation and 0.68 for motor curve formation. If the SR is greater than these cut-offs, consider (a) moving the solver points further toward the extremes of the stimulation space, if possible; (b) taking multiple trials at the already established point positions and trial-averaging the pulse parameters before re-solving for the Weiss equation; or (c) sampling more (*PW*, *PA*) points in between the solver points and performing a non-linear fit.

### Mechanistic insights and clinical relevance of stimulating at across the SD curve from modeling data

The modeling results presented here offer additional insights into how stimuli at different points along the SD curve vary. Specifically, high PA and high PW stimuli on the same SD curve activate many of the same axons yet systematically differ in the subsets of axons that are uniquely recruited. In the more horizontal regions of the curve (high PW), stimulation consistently activates axons closer to the electrode and, to reach the same intensity, recruits more small-diameter fibers. Conversely, in the more vertical regions of the curve (high PA), the uniquely activated axons are further away from the contact and include more large-diameter fibers (**Figure 8**). These results are consistent with previous modeling and experimental data that state that the difference in threshold of axon activation is the smallest between diameters at a high pulse width compared to low pulse width stimulation [58,4]. This effect is likely due to currents between nodes of Ranvier within the axon, which are greater in larger diameter axons [58]. Whereas previous studies have demonstrated the differences in axon activation in single axons across pulse widths [4,6], our results extend these findings to a full population of axons within a human-derived nerve model. By considering population-level averages, these outcomes become directly applicable to our experimental data, enhancing its explanatory power for both functional electrical stimulation (FES) and sensory restoration. Notably, the same systematic differences in activated axon populations appear in both motor and sensory data.

The models indicate that higher-intensity stimuli on the same SD curve recruit increasingly similar axon populations. DSC analysis with Kruskal-Wallis tests confirms that the lowest-intensity SD curves activate more distinct axon populations than those at higher intensities in both sensory and motor models. This suggests that alternating between stimuli along an SD curve at low intensities may produce more distinguishable sensations in sensory stimulation and recruit different motor units compared to high-intensity stimuli. Clinical validation is needed to determine how unique the two axon populations need to constitute a meaningful difference in activation. However, intraneural microstimulation research has shown that single sensory fiber activation is perceptible [59], supporting the clinical relevance of these findings to differentially modulate percepts.

These distinct axonal populations on either end of an SD curve have different implications for both sensory restoration and FES. In sensory restoration, the smaller-diameter fibers and more spatially contiguous regions of axons recruited by stimulation on the horizontal region of the SD curve could result in a smaller perceived area of sensation—even though the overall intensity may be similar to stimulation in the vertical region of the curve. Furthermore, activating large and small fibers together in a confined region likely alters the perceived quality of sensation. In FES for motor restoration, our models indicate that stimulation on the horizontal region of the SD curve more closely mimics the natural recruitment order by activating smaller fibers earlier. It also recruits more contiguous axonal regions, leading to more anatomically aligned muscle activation [60]. However, alternating between stimulation in the vertical and horizontal regions on a single contact could help reduce fatigue by engaging different motor units within the same muscles [10]. Therefore, the modeling results demonstrate practical reasons for stimulating at multiple points along an SD curve and explain why equally intense stimuli could still produce different outcomes.

### Using the SD curve to construct FES recruitment surfaces to improve selectivity, accuracy, and fatigue resistance of motor neuroprostheses

Because of the SD curve’s accuracy and robustness, it has tremendous potential for improving FES neuroprostheses and other motor neuron-targeting neuromodulation. We propose a novel recruitment surface construction method based on the SD curve’s ability to be predicted from just two distanced points. The proposed method acquires two sets of recruitment curves, a PW modulated set at the maximum PA value and PA modulated set at the maximum PW value. Each set would contain one recruitment curve for each muscle of interest activated by the stimulated nerve. These sets could be acquired efficiently by simultaneously recording from each muscle. SD curves would then be solved for between points with equal EMG values on the two recruitment curves across the full range of muscle activation for each muscle.

This novel characterization method would require an order of magnitude fewer points than a procedural sampling of the space and therefore make it practical to use the entire stimulation space rather than the few slices that recruitment curves have historically provided. Though not directly tested in this study, it is likely that our findings, and the expected benefits, generalize to many other types of motor neuron stimulation.

#### Inter-muscle selectivity for increased dexterity

For surface and implanted motor neuron stimulation, multiple muscles are often activated by a single electrode contact, and these relationships between muscles can covary in unpredictable ways. The *in silico* work of this study suggests that different populations of axons can be activated as PA and PW are varied. Full characterization of the stimulation space will maximize single contact muscle selectivity by utilizing the differences in muscle activation in the vertical and horizontal regions of the SD curve. This increased ability to more independently modulate individual muscles should allow for more accurate and dexterous movements.

#### Shallower slope for increased resolution of activation

It has long been theorized that recruitment curves with shallower slopes provide better resolution of muscle activation [12–15]. Shallower slopes provide more selectable stimulation values between minimum and maximum and therefore should provide greater resolution of the output. By characterizing the full stimulation space, it is possible to automatically find the shallowest path from threshold to maximum, even if that path requires PW and PA to be modulated independently and/or nonlinearly.

#### Intra-muscle selectivity for fatigue management

A single muscle is made up of many motor units, each activated by its own motor neuron. The number of motor units in the human extensor digitorum brevis for example has been estimated to be around 200 [61]. Therefore, as long as the muscle is not being activated maximally, multiple combinations of these motor units could be recruited to achieve an identical force from that muscle. Interleaved stimulation activating non-identical populations of motor units has been shown to decrease fatigue while maintaining activation strength in intramuscular [8], intrafascicular [9], and cuff electrodes [10]. Modulating PA and PW independently would make it easier to activate motor unit populations with less overlap, therefore increasing stamina.

### Using the SD curve for unique percept generation and improved modulation resolution in sensory neuroprostheses

#### Percept modulation via unique neural subset activation

Modeling data suggests that stimulating on the horizontal end of the SD curve (higher PW) tends to recruit smaller fibers closer to the cathodic contact, and stimulating on the vertical end of the curve (higher PA) tends to recruit larger fibers in a wider radius around the cathodic contact while leaving some of the closer, smaller fibers inactivated (**Figure 8**). Given that the activation of a single afferent fiber is perceivable [59,62,63], we would expect the differences in the fiber populations recruited at each end of an SD curve to be perceivable in sensation location or quality. The manipulation of these percept features independent of intensity would be highly impactful, as intensity covaries with these characteristics in standard PNS paradigms [27,64].

Regarding percept location, results were mixed in this study as to whether the reference stimulus in the horizontal region of the curve and trial stimuli in the vertical region elicited different sensations. Subject 1 reported sensation on the thumb and index finger for both sets of stimuli throughout the testing. However, for the curve just prior to muscle contraction, which the participant rated as mid-intensity in magnitude, a sensation on the intermediate phalanx of his third finger was elicited by the reference stimulus but diminished and eventually disappeared in the trial stimuli as PA increased. The participant also noted that for the percept on the middle finger to appear during the higher PA trial stimuli, he had to increase PW such that the percept on the thumb was significantly stronger than that elicited by the reference stimulus. Ultimately, Subject 1 decided to match the intensities on the thumb and index fingers between reference and trial stimuli to construct the curve. Subject 2, however, did not report any differences in percept location within a curve. Further study with a larger participant pool is needed to fully assess the potential for SD curves to modulate location while holding intensity constant.

With respect to percept quality, small temporal variations in stimulation paradigms have been shown to increase perceived naturalness [27]. In lieu of sinusoidal adjustments of a single pulse parameter, which affects intensity perception, stimulating at points positioned across an SD curve could maintain stimulus intensity while still varying neural population recruitment. Modulation within an SD curve also pairs well with high frequency stimulation paradigms that have the aim of eliciting more natural percepts by activating unique axon subsets [65,66].

#### Robustness to perceptual sensory adaptation

The absence of a significant difference in any sensory SD curve across three hours of data collection may indicate the robustness of SD curves to sensory adaptation (**Figure 7(b)**). All contacts showed evidence of perceptual adaptation, with an average increase in perceptual threshold of 0.075 mA at 250 µs, in addition to changes in estimated magnitude (**Figure 7(a)**). Applicably, SD curves may not need to be recalculated across several hours of use. Instead, to account for adaptation, a single point from each existing curve should be sampled to update the curve’s perceptual magnitude. New curves should only need to be generated at high intensities that were previously above the threshold for discomfort.

Mechanistically, the maintenance of each SD curve across the two-dimensional space may point to adaptation as a centrally mediated process rather than a change in peripheral nerve dynamics. This aligns with previous hypotheses on the nature of tactile temporal inhibition [67,68]. As demonstrated in the *in silico* data, intensity-matched stimuli at high PA and high PW tend to activate neural populations that contain a subset of unique fibers. While we have not yet proven that the same trends hold clinically beyond the anecdotal percept location changes from Subject 1, if adaptation was peripherally mediated, we would expect the reference stimulus in the horizontal region of the curve to adapt faster than the trial stimuli toward the vertical region of the curve due to the repeat activation of the reference stimulus. However, the absence of any shift in the generated curve demonstrates that adaptation is occurring equally to all subsets of that contact’s neural population. This points to adaptation occurring proximal not only to mechanoreceptors as previously shown [68] but to peripheral nerves as a whole. To better support this theory, further research is needed to prove that different neural populations are activated at different parts of the SD curve in a clinical model.

#### Investigating perceptual recruitment surfaces for multi-dimensional JNDs and increased modulation resolution

While recruitment curves and surfaces are standard in motor PNS research, they are less commonly applied in sensory PNS to map the entire dynamic intensity range due to the subjective nature of intensity perception as well as the time required to collect the data. The use of two-point SD curves to map the space drastically reduces the temporal burden of this methodology, and while perceived intensity is still a subjective output, the relationship between two points as described through psychophysics is not.

Understanding the shape of a recruitment surface can provide insight into multi-dimensional Just Noticeable Difference (JND) intensity measurements. Previous studies comparing the relative impact of different stimulation parameters on perceived intensity have chosen one or two points and evaluated the Weber fractions of each parameter at that point [26,39]. However, the relative effect of PA and PW is variable depending on where the selected point is on a strength-duration curve. Two-dimensional JNDs that examine the gradient of a perceptual sensory recruitment surface are a more accurate and applicable method of evaluating the impact of the two parameters on intensity.

Further, as similarly described for motor PNS, knowledge of the PA-PW surface can facilitate an increase in neural output resolution. Modulating both PA and PW enables the degree of change in perceived intensity to be controlled by modulating with respect to the gradient of the recruitment surface or the slope of the nearest SD curve. For example, adjusting stimulation nearly parallel to a SD curve will result in only a small change in perceived intensity compared to an equivalent stimulation adjustment oriented perpendicular to the same SD curve. By moving across a multi-dimensional space, discriminable differences in percept intensities that were previously within a single stimulator step size become accessible. Fully characterizing the two-dimensional charge space enables more evenly graded changes in stimulation, which is valuable for object recognition and compliance detection [64].

#### Circumventing technical limitations for increased evoked intensity range

Depending on participant physiology and interface specifications, a participant’s full dynamic range is sometimes not accessible when modulating in a limited parameter space. For example, some contacts on Subject 2’s median nerve C-FINE could not be used for this study because the charge limitation for the Shannon curve is below the charge required to evoke high-intensity sensations at long PWs [69]. Because of the nature of the charge-duration relationship, which describes how the charge required to elicit a neural response increases linearly with pulse width, this limitation can be mitigated by stimulating at higher PAs and correspondingly shorter PWs.

There can also be stimulation constraints at high PAs due to stimulator compliance voltage or current density restrictions. If an interface has high impedance, such as in transcutaneous stimulation, the compliance voltage of the stimulator may be reached at a lower PA than what is required to elicit a strong percept. Similarly, transcutaneous PNS may benefit from lower PA stimulation that decreases the current density administered at the skin to reduce discomfort and unwanted percepts caused by activation of local afferents in the skin [23,64,70,71]. This is particularly true for interfaces using current-controlled stimulators, in which high current density can cause burns due to changes in skin-electrode contact area [23,72]. Usually, decreasing current density requires increasing the electrode size, which in turn hampers stimulation selectivity [70]. Overall, parameter space characterization via SD curves enables neuromodulators to avoid these interface-based pitfalls by facilitating simultaneous PA and PW modulation nearer to the center of the charge space.

### SD curve applications in mixed motor and sensory stimulation

While PNS often focuses solely on either motor or sensory activation, most relevant peripheral nerves are mixed nerves containing both motor and sensory fibers. Thus, selectivity between motor and sensory activation is important for practical PNS neuroprostheses. For motor neuroprostheses, it is important to avoid painful or distracting sensory fiber activation while performing functional tasks. For sensory neuroprostheses, unintended muscle activation can affect EMG control of a prosthetic limb or elicit conflicting proprioceptive percepts. Unwanted muscle activation also limits the upper portion of the functional sensory dynamic range, as users experience uncomfortable muscle cramping at a lower current than their maximum comfortable sensory percept.

Our *in silico* results suggest that there are differences in the locations and sizes of the axon populations that are activated on either side of an SD curve. Given the difference in diameter sizes between motor and sensory fibers, it is likely that specific regions of the PA-PW stimulation space selectively favor motor and sensory activation. In theory, stimulation on the vertical side of the curve should preferentially activate motor fibers due to their larger diameters, while stimulation on the horizontal side of the curve should preferentially activate sensory fibers due to their smaller diameters. By acquiring two strength-duration curves, one that represents the threshold of activation for the desired modality and another that represents the maximum acceptable activation of the off-target modality, motor and sensory activation can be detangled. Using this method, the complete region of stimulation parameters that avoid unwanted motor or sensory activation can be quickly and easily defined.

Efficient characterization of the two-dimensional space also facilitates research into joint motor and sensory topics such as proprioception. For example, the effect of pulse width on the strength of the H-reflex with respect to the M-wave is an active area of research, and utilizing both motor and sensory SD curves can aid in the exploration of H-reflex activation while maintaining a constant M-wave [7,73,74].

### Limitations

#### Sample size

While the motor and sensory analyses in this study consist of 2,312 and 486 sampled points, respectively, the number of participants that the data were acquired from was limited. In the motor analysis, data were only acquired from a single participant with 5 nerve cuff electrodes (2 contacts on each) tested within that participant’s arm and shoulder. In the sensory analysis, data were acquired from two participants with different levels of experience with sensory stimulation studies. One nerve cuff electrode was tested each in Subject 1 (1 contact) and Subject 2 (3 contacts). This limited subject population is due to the rarity of the relevant injuries and the intensive nature of the implantation and research. While these results should be validated in more people, we posit that the findings will be generalizable given they align with the larger body of SD curve literature and across all 50 motor curves and 16 sensory curves acquired.

For the qualitative data collected from sensory participants such as magnitude estimation and percept location, the small sample size limits the strength of some of the proposed applications due to the differential experience between Subject 1 and Subject 2. Subject 1 has 8 years of experience with PNS studies and has accordingly developed a comparatively precise system for how he reports percepts. For example, Subject 1 commonly relates his estimated magnitudes to increases in percept areas, so he quickly noticed when different ends of the curve produced slightly different percept areas at equal intensities. Data collection for Subject 2 took place during his first year enrolled in the study, and as a result his system for reporting percepts was less consistent. Future studies with more participants are needed to further explore the application of SD curves to sensory percept manipulation.

#### Differences in target reference consistency between motor and sensory experiments

Even though no significant temporal shifts were detected in the sensory contour data, 60% of the equal muscle activation contours showed a significant difference (*p* < 0.05) between data collected in different experimental blocks. This likely results from differences in the target references between the two modalities. For the motor experiments, each target EMG value was determined at the beginning of the session as a percentage of the maximum EMG recorded for that muscle. This means that any changes in neural response or muscle activation that occurred over the course of data acquisition were not reflected in this target value. For the sensory experiments, the target reference was the live perceived intensity at a specific (*PW*, *PA*) point, meaning any changes in sensory perception were reflected in both the reference and sampled points. It is most likely that, in the same way that the sensory curves stayed consistent while the reported magnitude changed across blocks (**Figure 7**), the differences seen between blocks in the motor results could have been controlled for by renormalizing target EMG to an updated maximum EMG throughout the session.

#### Quantification of error in terms of PA rather than intensity

Throughout this paper, we quantify goodness of fit by evaluating error as the distance between the PA values our models predict at given PWs and the PA values of our samples at those PWs. Through this analysis, we see consistently high R^2^ values across many different conditions. However, the link between these high R^2^ values and the functional outcomes that could be obtained by applying our proposed fit methods to motor or sensory characterization is not entirely clear. Since muscle activation as a function of PA and/or PW has been repeatedly shown to be more sigmoidal than linear [20,21,75], different levels of muscle activation could have very different changes in EMG error for the same error in PA. This was seen via visual inspection of the acquired data, where SD curves that were in regions of the recruitment surfaces that had shallower slopes had lower R^2^ values than those in regions with steeper slopes. This aligns with the small but significant differences seen in the “any 2-point” fit analysis of activation level which showed higher R^2^ values towards the center of the activation range and lower values at the minima and maxima. This suggests that our analysis likely overestimates the ability of our curve fits to predict the actual outputs of interest in areas with steep slopes and underestimates the abilities in areas with shallow slopes. Similarly for sensory applications, the error in PA is not characterized here with respect to JNDs. Because intensity level affects JNDs per Weber’s law, and because sensory fiber recruitment also likely follows a sigmoidal shape as a function of charge [26], the precision in the PA-PW space required to remain within one JND is not necessarily consistent across all intensities.

## Conclusion

This work provides a clinical and *in silico* foundation for the application of strength-duration curves to characterize and improve outcomes of PNS for the control of muscle activation and restoration of somatosensation. The clinical data demonstrate that the strength-duration curve is an accurate model of muscle activation and sensory intensity contours across the full functional range of activation. Our analysis of different fitting techniques shows that solving the SD curve equation with the 2 points that are furthest into the horizontal and vertical regions of the curve provide significantly better curve fits than arbitrary combinations of 2 points. Further analysis shows that 2-point SD curve solutions with slope ratios under 0.68 for motor and 0.26 for sensory are within the expected distributions of the end-point solutions. The *in silico* modeling shows that one can expect to see greater activation of axons that are further away from the electrode and axons that are larger in diameter when stimulating in the vertical region of the SD curve as compared to the horizontal region.

The combination of these findings suggests many potential benefits of using the SD curve for the characterization of PNS and potentially other stimulation methodologies. For motor FES, characterizing the full stimulation space with SD curves could help create neuroprostheses that have better muscle selectivity, fatigue resistance, and fine motor control. For sensory PNS, SD curves could facilitate unique percept generation, agile adaptation adjustment, and high-resolution intensity modulation. For both modalities, the SD curve can be used to quickly define a comfortable stimulation region and avoid unwanted activation of the other modality. We also believe that this framework could have useful applications in other types of neurostimulation such as surface stimulation and spinal cord stimulation.

## Data Availability

All data produced in the present study are available upon reasonable request to the authors.

## Acknowledgements

We would like to thank the surgeons and clinical staff at University Hospitals and the Cleveland VA Medical Center for their clinical support and management of IRB compliance. We also thank W. Memberg and J. Dunning for engineering support, R. Kirsch for advice on motor data analysis, and E. Imka for graphic design. Last, we thank the participants and their families for their time and dedication to research improving outcomes for people with spinal cord injury and limb loss.

## Author Contributions

R.S.J. and B.J.A. contributed equally to experimental design, data collection, data analysis, and manuscript authorship. R.S.J. led the sensory work, and B.J.A. led the motor work. V.I.M. created the computational model and wrote the corresponding portion of the manuscript. A.B.A. advised on experimental design, result interpretation, and manuscript composition. D.J.T. conceived of the initial study idea and advised on experimental design, result interpretation, and manuscript composition.

